# Beyond IQ: Executive function deficits and their relation to functional, clinical, and neuroimaging outcomes in 3q29 deletion syndrome

**DOI:** 10.1101/2024.02.22.24303212

**Authors:** Rebecca M Pollak, Esra Sefik, Katrina Aberizk, Kuaikuai Duan, Roberto Espana, Ryan M Guest, Adam E Goldman-Yassen, Katrina Goines, Derek M Novacek, Celine A Saulnier, Cheryl Klaiman, Stormi Pulver, Joseph F Cubells, T Lindsey Burrell, Sarah Shultz, Elaine F Walker, Melissa M Murphy, Jennifer G Mulle

## Abstract

**Background:** 3q29 deletion syndrome (3q29del) is a rare (∼1:30,000) genomic disorder associated with a wide array of neurodevelopmental and psychiatric phenotypes. Prior work by our team identified clinically significant executive function deficits in 47% of individuals with 3q29del; however, the nuances of executive function in this population have not been described.

**Methods:** We used the Behavior Rating Inventory of Executive Function (BRIEF) to perform the first in-depth assessment of real-world executive functioning in a cohort of 32 individuals with 3q29del (62.50% male, mean age=14.50±8.26 years). High-resolution structural magnetic resonance imaging was performed on a subset of participants (n=24).

**Results:** We found global deficits in executive function; individuals with 3q29del scored significantly higher than the population mean on the BRIEF Global Executive Composite (GEC) and all subscales. 81.25% of study subjects (n=26) scored in the clinical range on at least one BRIEF subscale. BRIEF GEC T scores were significantly higher among 3q29del participants with a diagnosis of ADHD, and BRIEF GEC T scores were significantly associated with schizophrenia spectrum symptoms as measured by the SIPS. The BRIEF-2 ADHD Form accurately (sensitivity=86.70%) classified individuals with 3q29del based on ADHD diagnosis status, highlighting its potential use as a screener for ADHD in this population. BRIEF GEC T scores were significantly correlated with cerebellar white matter and subregional cerebellar cortex volumes.

**Conclusions:** Together, these data expand our understanding of the phenotypic spectrum of 3q29del and identify executive function as a core feature linked to both psychiatric and neuroanatomical features of the syndrome.

## Introduction

3q29 deletion syndrome (3q29del) is a rare (1:30,000) (Kendall et al., 2017; Stefansson et al., 2014) genomic disorder caused by the 1.6 Mb recurrent, typically *de novo* 3q29 deletion (hg19, chr3:195725000-197350000) (Ballif et al., 2008; Glassford et al., 2016; Willatt et al., 2005). The clinical phenotype of 3q29del is heterogeneous, ranging from mild to moderate intellectual disability (ID) (Ballif et al., 2008; Cox & Butler, 2015; Glassford et al., 2016; Klaiman et al., 2022; Sanchez Russo et al., 2021; Willatt et al., 2005) to a 19-fold increased risk for autism spectrum disorder (ASD) (Itsara et al., 2009; Pollak et al., 2019; Sanders et al., 2015) and a greater than 40-fold increased risk for schizophrenia spectrum disorders (SZ) (Kirov et al., 2012; Marshall et al., 2017; Mulle, 2015; Mulle et al., 2010; Szatkiewicz et al., 2014). Recent work by our team has uncovered additional phenotypes associated with 3q29del, including attention deficit/hyperactivity disorder (ADHD) and graphomotor weakness (Klaiman et al., 2022; Pollak et al., 2023b; Sanchez Russo et al., 2021). Further, 47% of study participants were found to have clinically significant executive function deficits (Klaiman et al., 2022; Sanchez Russo et al., 2021). However, our understanding of the nuances of these phenotypes, including executive function, are still evolving.

Executive function refers to the constellation of higher-order cognitive processes that control and coordinate purposeful goal-directed behaviors (Baggetta & Alexander, 2016; Best & Miller, 2010; Miller & Wallis, 2009). It is generally agreed that executive function is comprised of three core components: inhibitory control, working memory, and cognitive flexibility (Baggetta & Alexander, 2016; Best & Miller, 2010; Miller & Wallis, 2009). These foundational functions are moderately inter-correlated yet separable, they can be differentially affected in different patient populations, and they serve as the basis for more complex cognitive constructs, such as planning, problem solving and abstract reasoning (Diamond, 2013; Miyake et al., 2000). Executive function allows an individual to integrate information about their goals with sensory input to guide actions in an adaptive and dynamic manner, in accordance with the demands of the present context. Executive function deficits can have an adverse impact on academic and occupational achievement, mood regulation, and social function (Baggetta & Alexander, 2016; Best & Miller, 2010; Miller & Wallis, 2009). Studies have identified executive function deficits in individuals with ADHD (Biederman et al., 2004; Brown, 2009; Marije Boonstra, Oosterlaan, Sergeant, & Buitelaar, 2005; Martel, Nikolas, & Nigg, 2007) and SZ (Kraepelin, 1913; Lysaker et al., 2008; Orellana & Slachevsky, 2013; Pickup, 2008; Velligan & Bow-Thomas, 1999; Wobrock et al., 2009), as well as genomic disorders with phenotypic similarities to 3q29del, including 22q11.2 deletion syndrome (Albert, Abu-Ramadan, Kates, Fremont, & Antshel, 2018; Everaert et al., 2023; Gur et al., 2023; O’Hora et al., 2023). Notably, executive function ability has been shown to associate with later-onset phenotypes in children with 22q11.2 deletion syndrome (Albert et al., 2018), highlighting the importance of understanding executive function and its links to neurodevelopmental and psychiatric phenotypes, as well as its potential as an early treatment target with promising benefits for both childhood and adult outcomes.

The prefrontal cortex has canonically been associated with executive function; however, it is now clear that these processes rely on distributed neural networks and emerging evidence points to a pivotal role for the cerebellum. Convergent evidence from neuropsychological testing, neuroimaging, humans with focal brain damage, and non-human animal studies have identified the cerebellum as a critical brain region for higher-order cognitive processing, including executive function (Bellebaum & Daum, 2007; Deverett, Koay, Oostland, & Wang, 2018; Koziol, Budding, & Chidekel, 2012; Schmahmann, 2019; Schmahmann & Sherman, 1998). Notably, neuroimaging studies by our team have identified a particularly high frequency of structural anomalies in the posterior fossa of individuals with 3q29del, surpassing the combined occurrence of radiological anomalies found in all other brain regions; for example, more than 60% of 3q29del study participants have cerebellar hypoplasia and/or cystic or cyst-like malformations around the cerebellum, and both cerebellar cortex and white matter show significant volumetric differences compared to typically developing controls (Sanchez Russo et al., 2021; Sefik et al., 2022). These cerebellar abnormalities, coupled with the increased rate of executive function deficits identified in individuals with 3q29del, highlight the importance of understanding this complex cognitive phenotype and its relationship with neurodevelopmental and psychiatric morbidity in 3q29del.

The present study is the first detailed description of executive function abilities assessed in individuals with 3q29del. We define the profile of executive function and we explore the relationship between executive function and general cognitive ability, neurodevelopmental and psychiatric phenotypes, and cerebellar volume. This study is an important contribution to our evolving understanding of 3q29del; executive function sits at the nexus of multiple neurodevelopmental and psychiatric phenotypes associated with 3q29del and may help to explain some of the underlying mechanisms contributing to the substantial psychiatric comorbidity experienced by this population. The results from this study will help to guide future research to further explore executive function in this population, as well as targeted interventions to improve executive function abilities in individuals with 3q29del.

## Methods

See Supplemental Information for detailed Methods.

### Study participants

Individuals with 3q29del were recruited from the online 3q29 registry (3q29deletion.org) for 2 days of in-person deep phenotyping, as previously described (Klaiman et al., 2022; Murphy et al., 2018; Sanchez Russo et al., 2021). Informed consent was provided by all participants over 18 years of age; for participants under 18 years of age, a parent or guardian provided informed consent and the study participant provided informed assent. 32 individuals with 3q29del (62.5% male) were included in the present study. Study participants ranged in age from 4.85 to 39.12 years (mean=14.50±8.26 years). See Table 1 for a description of the study sample. This study was approved by Emory University’s Institutional Review Board (IRB00064133) and Rutgers University’s Institutional Review Board (PRO2021001360).

**Table 1.** Demographic information for study participants with 3q29 deletion syndrome (n=32).

| | | Mean ± SD | Range |
| --- | --- | --- | --- |
| Age (years) | | 14.50 ± 8.26 | 4.85–39.12 |
| Composite IQ | | 73.03 ± 14.18 | 40–99 |
|  |  | **N** | **%** |
| Sex |  |  |  |
|  | Male | 20 | 62.50% |
|  | Female | 12 | 37.50% |
| Race |  |  |  |
|  | White | 29 | 90.63% |
|  | More than one race | 3 | 9.37% |
| Ethnicity |  |  |  |
|  | Hispanic/Latino | 1 | 3.13% |
|  | Not Hispanic/Latino | 31 | 96.87% |

### Measures

The measures used in this study were as previously described (Klaiman et al., 2022; Murphy et al., 2018; Sanchez Russo et al., 2021).

### Neuroimaging

High-resolution structural magnetic resonance imaging (MRI) data were collected and processed as previously described (Sanchez Russo et al., 2021; Sefik et al., 2022).

### Analysis

All analyses were performed in R version 4.0.4 (R Core Team, 2008). Due to the small sample size, most analyses were considered exploratory and unadjusted p values were reported. Statistical analysis was performed using simple linear models and one-sided, one-sample Student’s t tests implemented using the stats R package (R Core Team, 2008). P values for simple linear models were calculated using robust standard errors via the sandwich and lmtest R packages (Hothorn et al., 2015; Zeileis, Lumley, Berger, Graham, & Zeileis, 2019). All models were adjusted for age and sex. For neuroimaging data, multiple linear regression models were constructed adjusting for age and sex; for models where there was a significant relationship with the absolute brain region volume, models were re-run adjusting for age, sex, and estimated total intracranial volume (eICV) to test for regional specificity beyond the influence of global variability in head size. Receiver operating characteristic (ROC) curves were constructed using the ROCit R package (Khan & Brandenburger, 2020). Data visualization was performed using the plotly R package (Sievert et al., 2017).

## Results

### Global executive function deficits in 3q29 deletion syndrome

Higher scores on the BRIEF indicate worse executive function; a T score of 70 or higher indicates a clinically significant deficit. On average, study participants with 3q29del scored significantly higher than the population mean T score of 50 on the Global Executive Composite (GEC) (mean=67.81±10.68, p < 0.001) and across all subscales (all p < 0.001, Figure 1A, Table S2). There were no significant differences between males and females with 3q29del across all domains of the BRIEF (Figure S1). 15 study participants with 3q29del (46.88%) scored above the clinical threshold of 70 on the GEC, indicating clinically significant executive function deficits. Of the nine subscales, study participants with 3q29del showed the most impairment on the Shift subscale (mean=69.16±12.52), and the least impairment on the Organization of Materials subscale (mean=60.09±11.65, (Figure 1A, Table S2). Study participants with 3q29del demonstrated a range of impairments across BRIEF subscales; six participants (18.75%) did not score above the clinical threshold on any subscales, while two participants (6.25%) scored above the clinical threshold on all nine subscales (Figure 1B). A majority of study participants with 3q29del (n=22, 68.75%) scored above the clinical threshold on two or more subscales and 81.25% of study participants with 3q29del (n=26) scored above the clinical threshold on at least one subscale (Figure 1B), further emphasizing the substantial burden of executive dysfunction in this population. Together, these data demonstrate significant adverse impacts to executive function abilities in individuals with 3q29del.

**Figure 1.**
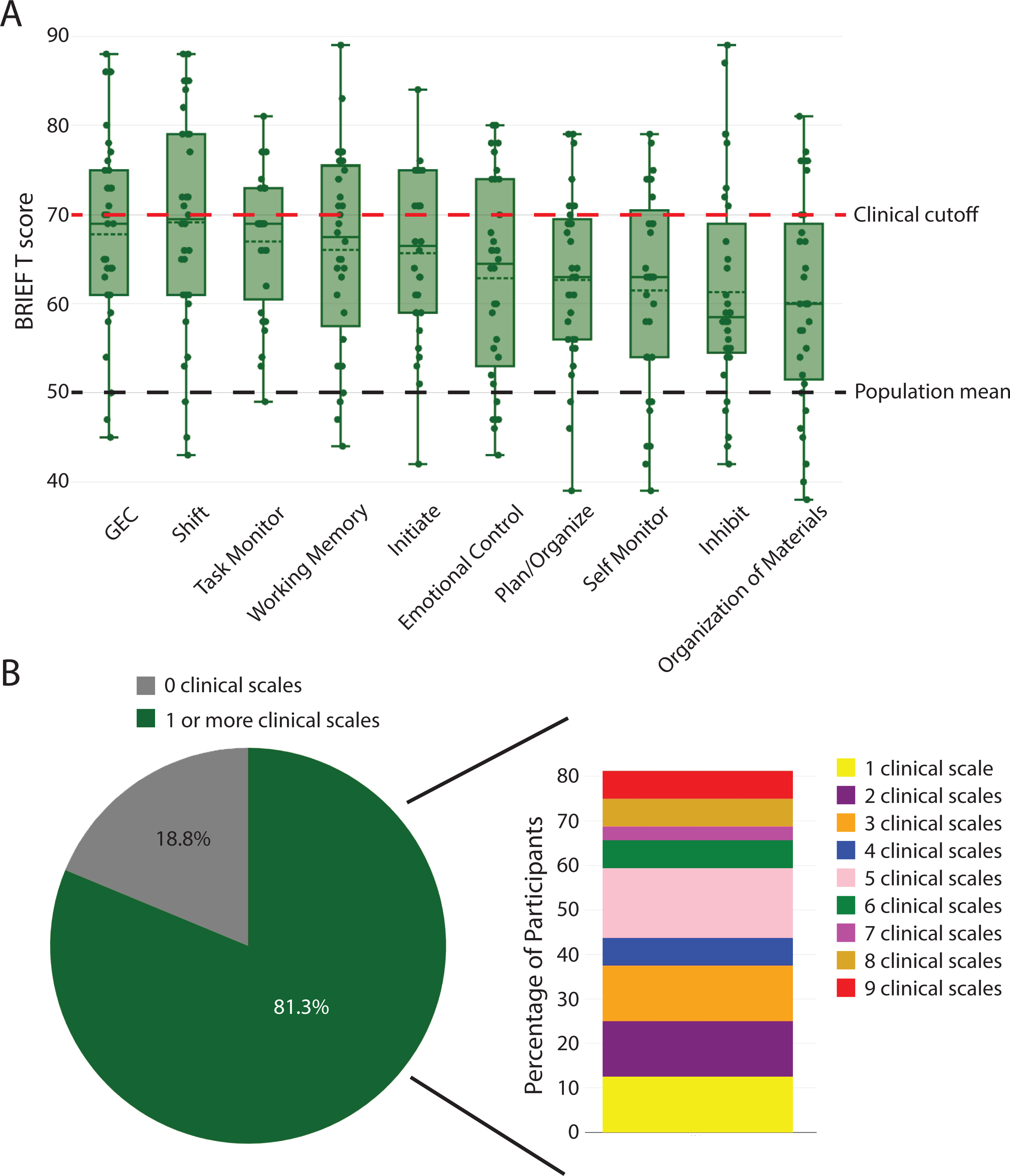
**A)** Distribution of T scores on the BRIEF GEC and BRIEF subscales for study participants with 3q29del (n=32). The black dashed line indicates the population mean; the red dashed line indicates the clinical cutoff. Subscales are ordered left to right by decreasing mean severity. **B)** Pie chart showing the proportion of study participants with 3q29del (n=32) scoring in the clinical range (T scores ≥ 70) on one or more BRIEF scales, expanded to show the proportion of participants scoring in the clinical range on 1 to 9 BRIEF scales. 3q29del, 3q29 deletion syndrome; BRIEF, Behavior Rating Inventory of Executive Function; GEC, Global Executive Composite

### Executive function is orthogonal to cognitive ability in 3q29 deletion syndrome

The 3q29 deletion is commonly associated with mild to moderate ID (Ballif et al., 2008; Cox & Butler, 2015; Glassford et al., 2016; Willatt et al., 2005); in the present study, the mean composite IQ in study participants with 3q29del was 73.03±14.18, as previously reported (Klaiman et al., 2022; Sanchez Russo et al., 2021). However, there was substantial variability in IQ across study participants, ranging from moderate ID to normal cognitive ability (range=40-99) (Klaiman et al., 2022; Sanchez Russo et al., 2021). We sought to determine whether variation in executive functioning is correlated with variability in cognitive ability in our study participants. There was no significant relationship between BRIEF GEC T scores and composite (r^2^=-0.011, p=0.424), nonverbal (r^2^=0.038, p=0.146), or verbal IQ (r^2^=-0.033, p=0.920; Figure 2A-C). Together, these data demonstrate that executive function is orthogonal to cognitive ability in individuals with 3q29del, and that poor performance on the BRIEF is not an artifact of diminished cognitive ability in our study population.

**Figure 2.**
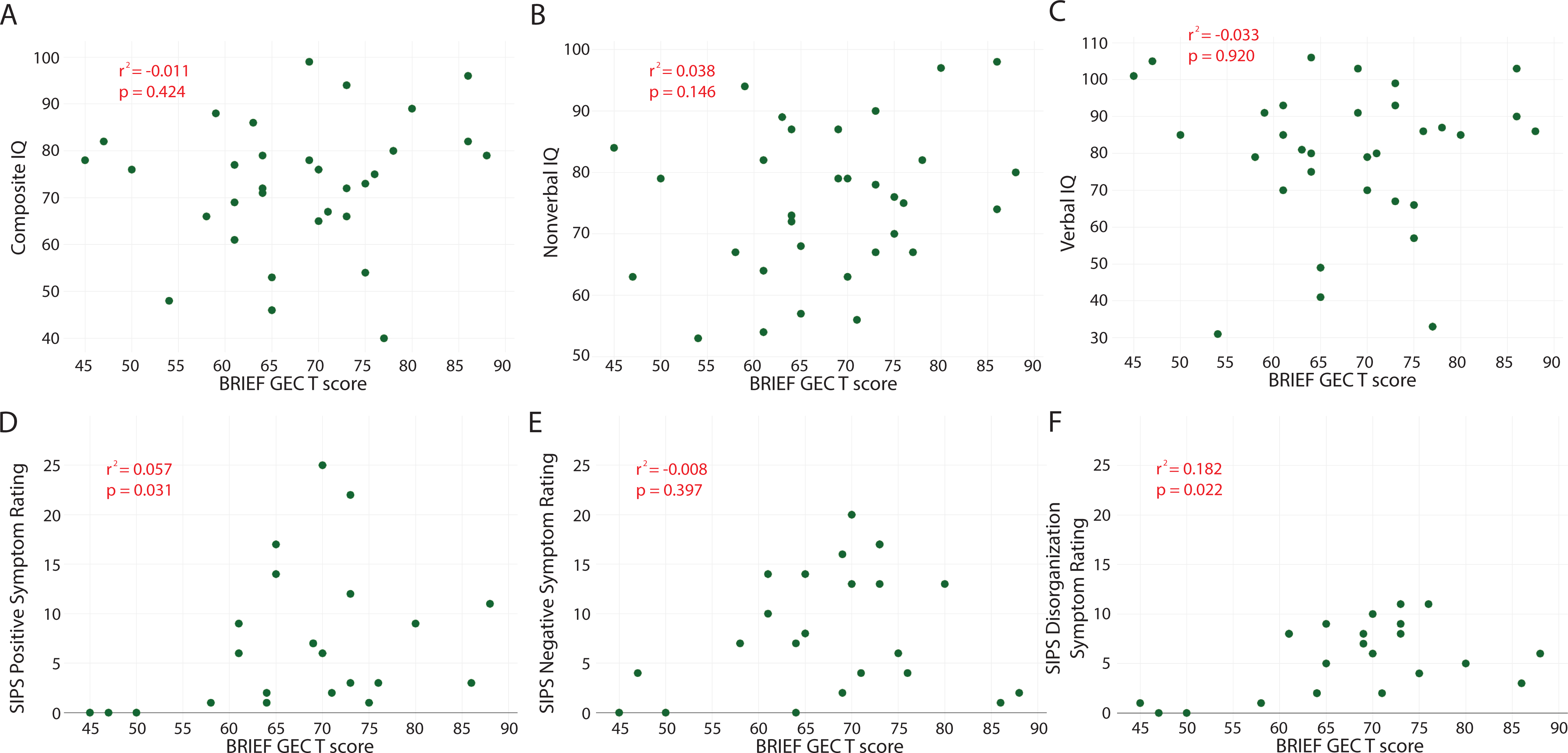
**A)** Correlation between BRIEF GEC T scores and composite IQ for study participants with 3q29del (n=32). **B)** Correlation between BRIEF GEC T scores and nonverbal IQ for study participants with 3q29del (n=32). **C)** Correlation between BRIEF GEC T scores and verbal IQ for study participants with 3q29del (n=32). **D)** Correlation between BRIEF GEC T scores and SIPS Positive Symptom Ratings for study participants with 3q29del (n=23). **E)** Correlation between BRIEF GEC T scores and SIPS Negative Symptom Ratings for study participants with 3q29del (n=23). **F)** Correlation between BRIEF GEC T scores and SIPS Disorganization Symptom Ratings for study participants with 3q29del (n=23). 3q29del, 3q29 deletion syndrome; BRIEF, Behavior Rating Inventory of Executive Function; GEC, Global Executive Composite; SIPS, Structured Interview for Psychosis-Risk Syndromes

### Executive function is correlated with psychosis spectrum symptoms

The 3q29 deletion is the largest known genetic risk factor for SZ (Mulle, 2015; Mulle et al., 2010; Singh et al., 2022); executive function deficits have a well-established association with SZ (Kraepelin, 1913; Lysaker et al., 2008; Orellana & Slachevsky, 2013; Pickup, 2008; Velligan & Bow-Thomas, 1999; Wobrock et al., 2009). We sought to define the relationship between executive function and psychosis spectrum symptoms in our study population. We found significant positive correlations between the BRIEF GEC and positive and disorganization symptoms endorsed on the SIPS (Figure 2D-F), indicating that individuals with 3q29del and poorer executive function experience more severe psychosis spectrum symptoms on average. The strongest correlation was between the BRIEF GEC and the SIPS disorganization symptom dimension (r^2^=0.182, p=0.022; Figure 2F). There was a weak relationship between the BRIEF GEC and the SIPS positive symptom dimension (r^2^=0.057, p=0.031; Figure 2D). There was no relationship between the BRIEF GEC and the SIPS negative symptom dimension (r^2^=-0.008, p=0.397; Figure 2E). These data show that executive function deficits are significantly associated with the severity of two major dimensions of psychosis spectrum symptoms in individuals with 3q29del.

### Psychiatric and neurodevelopmental comorbidity is associated with executive function deficits

Individuals with 3q29del are at increased liability for a wide range of neurodevelopmental and psychiatric phenotypes, including ASD, anxiety disorder, ADHD, and SZ (Ballif et al., 2008; Cox & Butler, 2015; Glassford et al., 2016; Itsara et al., 2009; Kirov et al., 2012; Klaiman et al., 2022; Marshall et al., 2017; Mulle, 2015; Mulle et al., 2010; Pollak et al., 2019; Sanchez Russo et al., 2021; Sanders et al., 2015; Szatkiewicz et al., 2014; Willatt et al., 2005). In the general population, ADHD and SZ are both associated with poorer executive function (Biederman et al., 2004; Brown, 2009; Kraepelin, 1913; Lysaker et al., 2008; Marije Boonstra et al., 2005; Martel et al., 2007; Orellana & Slachevsky, 2013; Pickup, 2008; Velligan & Bow-Thomas, 1999; Wobrock et al., 2009). To determine whether any individual neurodevelopmental or psychiatric diagnosis was associated with executive function in our study population, we compared the BRIEF GEC T scores for individuals with 3q29del with and without ASD, SZ prodrome/psychosis, ADHD, anxiety disorders, graphomotor weakness, ID, and enuresis (Figure 3A). We found that individuals with 3q29del and a diagnosis of ADHD have significantly worse executive function relative to individuals with 3q29del without ADHD (ADHD mean=72.80±8.06, no ADHD mean=59.50±9.42, p=3.08E-8; Figure 3A); there were no other significant relationships. These data demonstrate that the relationship between executive function and ADHD phenotypes is present in individuals with 3q29del.

**Figure 3.**
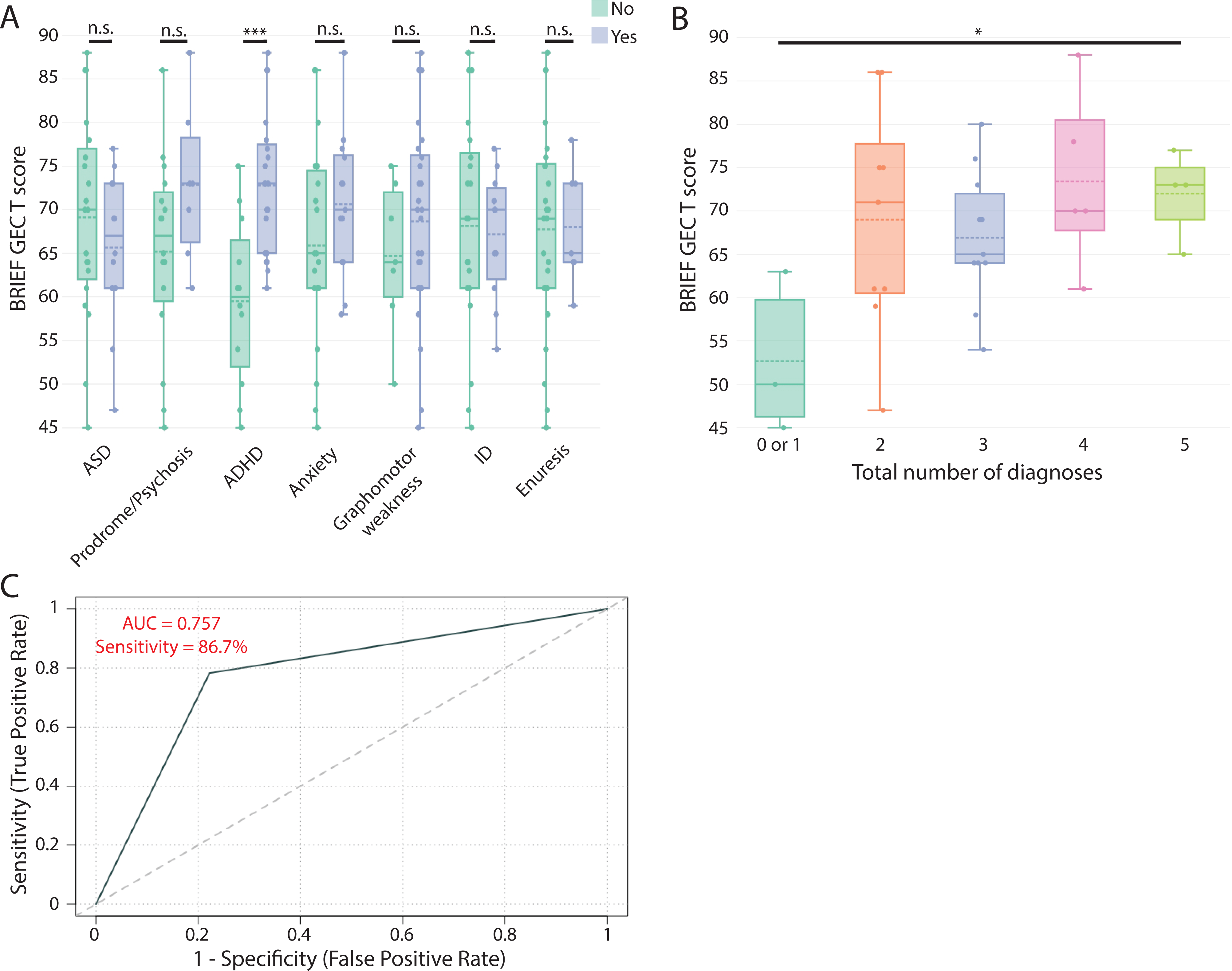
**A)** Distribution of BRIEF GEC T scores for study participants with 3q29del (n=32) with and without specific neurodevelopmental or psychiatric diagnoses. **B)** Distribution of BRIEF GEC T scores for study participants with 3q29del (n=32) with an increasing number of comorbid neurodevelopmental and psychiatric diagnoses. **C)** Receiver operating characteristic curve showing the ability of the BRIEF-2 ADHD Form to correctly classify study participants with 3q29del (n=26) with and without a diagnosis of ADHD. 3q29del, 3q29 deletion syndrome; BRIEF, Behavior Rating Inventory of Executive Function; GEC, Global Executive Composite; ADHD, attention-deficit/hyperactivity disorder

Prior work by our team has identified neurodevelopmental and psychiatric comorbidity as a hallmark feature of 3q29del (Pollak et al., 2023a; Sanchez Russo et al., 2021); we found that increasing degrees of comorbidity, rather than individual neurodevelopmental or psychiatric diagnoses, is significantly associated with poorer adaptive function in this population (Pollak et al., 2023a). We sought to determine whether there is a similar relationship between executive function and comorbidity in our study population. We found that BRIEF GEC T scores are correlated with comorbidity, where increasing comorbidity is significantly associated with poorer executive function (p=0.039; Figure 3B). Together, these data emphasize the central nature of neurodevelopmental and psychiatric comorbidity to the 3q29del phenotype, and show that comorbidity, rather than individual diagnoses, has a stronger relationship with executive function in individuals with 3q29del.

### The BRIEF-2 is an accurate screener for ADHD in individuals with 3q29 deletion syndrome

Screening tools are valuable instruments that can be used to prioritize individuals for diagnostic evaluations or to identify sub-populations for future studies. In the case of 3q29del, identifying effective screening tools for specific neurodevelopmental or psychiatric phenotypes will help to ensure that the highest-risk individuals will receive critical diagnostic evaluations as early as possible, while simultaneously reducing the burden of multi-disorder diagnostic batteries on caregivers and the healthcare system. Our team has identified ADHD as one of the most common psychiatric diagnoses in individuals with 3q29del, with 63% of individuals with 3q29del qualifying for a diagnosis of ADHD in a recent study (Sanchez Russo et al., 2021). ADHD is intimately linked to executive function (Biederman et al., 2004; Brown, 2009; Marije Boonstra et al., 2005; Martel et al., 2007); here, we sought to determine whether the BRIEF-2 ADHD Form is an accurate screening tool for ADHD in children with 3q29del (n=26). We found that the BRIEF-2 ADHD form had a sensitivity rate of 86.7%, indicating that 13.33% of individuals with ADHD did *not* screen positive on the ADHD Form (n=2), and a specificity rate of 60.00%, indicating that 40.00% of individuals that screened positive did not have an ADHD diagnosis (n=4) (Figure 3C). The BRIEF-2 ADHD form is a more sensitive screener for ADHD in individuals with 3q29del than the Achenbach Child Behavior Checklist DSM-keyed Attention-deficit/hyperactivity problems scale (Pollak, Mortillo, Murphy, & Mulle, 2024), which had a sensitivity of 72.2% and a specificity of 81.8. Together, these data show that the BRIEF-2 ADHD Form can be a useful screening tool for ADHD in children with 3q29del.

### Executive function ability is associated with cerebellar volumetric changes

While the prefrontal cortex has long been considered the locus of executive function, emerging evidence highlights the cerebellum’s contribution to higher-order cognition (Bellebaum & Daum, 2007; Koziol et al., 2012; Schmahmann, 2019; Schmahmann & Sherman, 1998). Work by our team has identified significant cerebellar abnormalities associated with 3q29del; for example, over 60% of 3q29del individuals show cerebellar hypoplasia (Sanchez Russo et al., 2021; Sefik et al., 2022). We sought to determine whether cerebellar anomalies are associated with executive function abilities in individuals with 3q29del. There was no relationship between BRIEF GEC T scores and total cerebellar volume (r^2^=-0.050, p=0.820; Figure 4B). There was a significant relationship between BRIEF GEC T scores and cerebellar white matter volume, where worse executive function is associated with *increased* cerebellar white matter volume (r^2^=0.349, p=8.11E-4; Figure 4C); this relationship persisted after adjusting for estimated intracranial volume (eICV) (r^2^=0.354, p=7.72E-4). There was a significant relationship between BRIEF GEC T scores and cerebellar cortical volume, where worse executive function is associated with *decreased* cerebellar cortical volume (r^2^=0.050, p=0.040; Figure 4D); however, this relationship did not persist after adjusting for eICV (r^2^=0.019, p=0.129). In a prior study our team identified an increased prevalence of posterior fossa arachnoid cysts and mega cisterna magna (PFAC/MCM) in individuals with 3q29del (Sanchez Russo et al., 2021; Sefik et al., 2022); however, BRIEF GEC T scores were not significantly different between individuals with 3q29del with and without PFAC/MCM (PFAC/MCM mean=66.23±9.19, no PFAC/MCM mean=67.09±11.77, p=0.650; Figure S2A).

**Figure 4.**
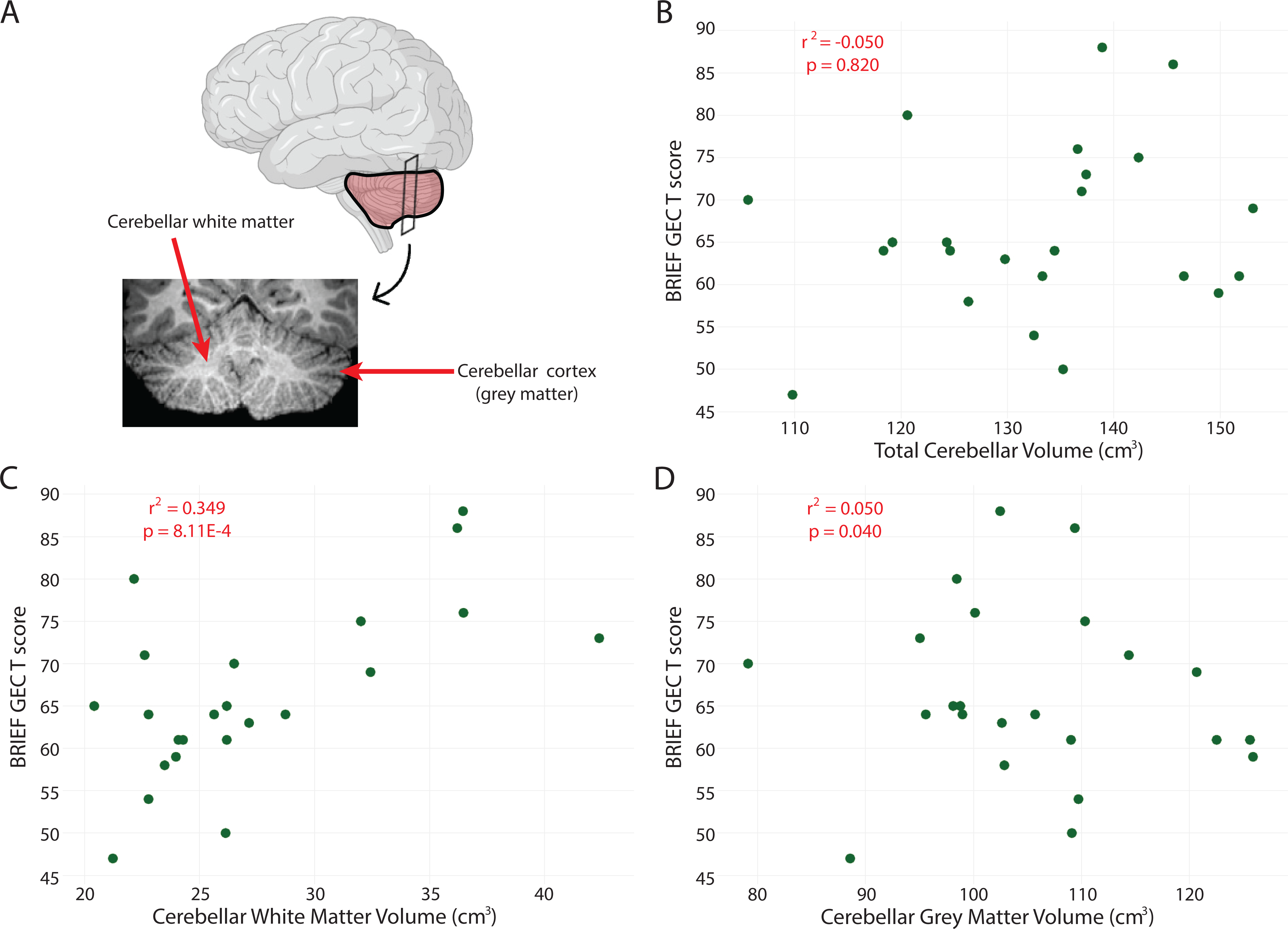
**A)** Diagram illustrating the cerebellum with a representative coronal image from a T1-weighted MRI showing cerebellar white matter and cerebellar cortex. **B)** Correlation between BRIEF GEC T scores and total cerebellar volume for study participants with 3q29del (n=23). **C)** Correlation between BRIEF GEC T scores and cerebellar white matter volume for study participants with 3q29del (n=23). **D)** Correlation between BRIEF GEC T scores and cerebellar cortical (grey matter) volume for study participants with 3q29del (n=23). 3q29del, 3q29 deletion syndrome; BRIEF, Behavior Rating Inventory of Executive Function; GEC, Global Executive Composite

To further explore the putative relationship between executive function and cerebellar cortical volume in study subjects with 3q29del, we analyzed the relationship between BRIEF GEC T scores and cerebellar cortex subregional volumes (Figure S2B-R). There were no significant relationships between BRIEF GEC T scores and any subregions of the anterior lobe of the cerebellar cortex (Figure S2C-D). Within the vermis, posterior lobe, and flocculonodular lobe of the cerebellar cortex, specific subregions were significantly associated with BRIEF GEC T scores. In general, increased volume of the subregion was associated with worse executive function. The following regions showed significant relationships with BRIEF GEC T scores that persisted after adjusting for eICV: left cerebellar hemisphere lobule VI volume (unadjusted r^2^=0.072, p=0.037; adjusted r^2^=0.134, p=0.038; Figure S2F); right cerebellar hemisphere lobule VIII volume (unadjusted r^2^=0.128, p=0.038; adjusted r^2^=0.154, p=0.037; Figure S2N); and left cerebellar hemisphere lobule IX volume (unadjusted r^2^=0.116, p=0.032; adjusted r^2^=0.153, p=0.026; Figure S2O). There were two regions that had a significant relationship with BRIEF GEC T scores that did not persist after adjusting for eICV: cerebellar vermis VI-VII volume (unadjusted r^2^=0.124, p=0.046; adjusted r^2^=0.154, p=0.056; Figure S2E); and right cerebellar hemisphere lobule X volume (unadjusted r^2^=0.174, p=0.045; adjusted r^2^=0.190, p=0.062; Figure S2R). Together, these data demonstrate a significant relationship between executive function and cerebellar white matter volume in individuals with 3q29del and reveal subregion-specific relationships between executive function and cerebellar cortical volume localized mostly to the posterior lobe of the cerebellar cortex, consistent with the presence of a functional topography within the cerebellum (Catherine J. Stoodley, Desmond, Guell, & Schmahmann, 2020; C. J. Stoodley & Schmahmann, 2010).

## Discussion

The present study is the first detailed description of executive function abilities in individuals with 3q29del. We identified global deficits in executive function, with significantly elevated mean scores on the BRIEF GEC as well as across all nine BRIEF subscales. The Shift subscale, a measure of cognitive flexibility, showed the highest mean impairment in 3q29del. Individuals with 3q29del and a diagnosis of ADHD had significantly poorer executive function relative to individuals with 3q29del without ADHD; poorer executive function was also significantly associated with more severe SZ spectrum symptoms as measured by the SIPS. Additionally, the BRIEF-2 ADHD Form accurately discriminated between individuals with 3q29del with and without a diagnosis of ADHD, highlighting its potential role as a time-efficient screening tool in this population. Neurodevelopmental and psychiatric comorbidity was associated with executive function, with an increasing number of diagnoses corresponding to significantly poorer executive function. Furthermore, BRIEF GEC scores showed no correlation with cognitive ability, indicating that mechanisms underlying cognitive ability and everyday executive functioning may be orthogonal in 3q29del, while both factors contribute to adaptive behavior as reported previously (Pollak et al., 2023a). Finally, executive function was significantly correlated with volumetric measures of the cerebellum, potentially identifying neuroanatomical changes contributing to executive function deficits in this population.

While this is the first description of executive function in 3q29del, deficits in executive function in this population are not without precedent. Genomic disorders with phenotypic similarities to 3q29del, including 22q11.2 deletion syndrome (Albert et al., 2018; Everaert et al., 2023; Gur et al., 2023; O’Hora et al., 2023), have documented evidence of significant executive function deficits. There is a large body of literature surrounding executive function abilities in 22q11.2 deletion syndrome specifically; executive function deficits in this population have been identified across the lifespan, from preschool-aged children to adults (Albert et al., 2018; Everaert et al., 2023; Gur et al., 2023; O’Hora et al., 2023). Worsening executive function is associated with psychosis spectrum symptoms in children with 22q11.2 deletion syndrome (Gur et al., 2023), and measures of childhood executive function can predict young adult outcomes in this population, including symptoms of psychosis (Albert et al., 2018). These data are similar to our findings in the present study of individuals with 3q29del and suggest that executive function may be a core feature central to understanding the wide neurodevelopmental and psychiatric phenotypic spectrum associated with the 3q29 deletion, including the substantial risk for conversion to psychosis.

In addition to the links between executive function and genomic disorders phenotypically similar to 3q29del, executive function deficits have also been long associated with idiopathic SZ and ADHD. Indeed, concepts related to executive function have been associated with the SZ phenotype since the early 20^th^ century (Kraepelin, 1913); executive function deficits are the most common cognitive phenotype in individuals with SZ (Orellana & Slachevsky, 2013; Velligan & Bow-Thomas, 1999; Wobrock et al., 2009). Executive function deficits are also a core feature of ADHD (Brown, 2009) and are present across the lifespan (Biederman et al., 2004; Marije Boonstra et al., 2005; Martel et al., 2007). We identified a relationship between executive function and diagnoses of ADHD, as well as SZ spectrum symptom severity, in our study sample of individuals with 3q29del. The concordance between our findings and results from studies of idiopathic cases of SZ and ADHD suggest that understanding executive function deficits in the context of the 3q29 deletion may provide generalizable insights to the study of SZ and ADHD at large.

Cognitive ability, specifically composite IQ, has canonically been used as a proxy for an individual’s level of everyday functioning. The results of the present study, notably the astonishingly high rate of clinically significant executive function deficits alongside the lack of a relationship between cognitive ability and executive function, suggest that IQ alone may not be a sufficient measure to understand real-world function for individuals with 3q29del. Traditional cognitive testing may overlook impairments in crucial aspects of higher-order cognitive functioning that are relevant to everyday behaviors in this population. This finding is aligned with prior reports that have shown that certain executive functions, including cognitive flexibility, are either uncorrelated with or are relatively weakly related to IQ (Ardila, Pineda, & Rosselli, 2000; Friedman et al., 2006), suggesting that traditional intelligence tests are insufficient in gauging the full spectrum of fundamental executive control abilities essential for various behaviors. This is of particular concern for individuals with 3q29 deletion syndrome who have IQ scores well within the normal range alongside compromised executive function; IQ scores in the normal range may create a perception of academic ability that is not accurate, given the co-occurring executive function deficits. Together, these data suggest that individuals with 3q29del should be clinically evaluated using measures of both cognitive ability and executive function as a standard of care. Furthermore, plans for treatment, management, and educational and occupational support should explicitly address and support executive function deficits.

We identified a significant relationship between cerebellar volumetric measures and executive function in the present study. There is emerging evidence linking the cerebellum to a range of cognitive processes, including executive function (Bellebaum & Daum, 2007; Koziol et al., 2012; Schmahmann, 2019; Schmahmann & Sherman, 1998). Changes in cerebellar structure and function have been identified in children with ADHD (Bechtel et al., 2009; Tomasi & Volkow, 2012), suggesting that cerebellar anomalies may contribute to the pathogenesis of ADHD. The cerebellum is also thought to have a role in SZ, due to its role in cognitive processes and the increased incidence of cerebellar anomalies in individuals with SZ (Andreasen & Pierson, 2008; Picard, Amado, Mouchet-Mages, Olié, & Krebs, 2008; Yeganeh-Doost, Gruber, Falkai, & Schmitt, 2011). Consistent with the findings of the present study, increased cerebellar white matter volume has been reported in idiopathic SZ, possibly indicating abnormal cerebellar connectivity (Lee et al., 2007). Our findings suggest that this is another point of convergence between 3q29del and idiopathic SZ; further exploration of cerebellar connectivity and executive function in 3q29del may shed light on the neuroanatomical underpinnings of these complex phenotypes. In an exploratory analysis, we also identified significant relationships between executive function and cerebellar cortical subregional volumes, where increased subregional volume was associated with poorer executive function. Future studies are required to replicate these exciting preliminary findings and may yield additional insight into altered cerebellar connectivity in 3q29del.

Executive function deficits can have a major impact on day-to-day function, but there are techniques and interventions that can improve an individual’s executive function abilities. Targeted interventions to improve executive function can be applied across the lifespan; studies have shown efficacy in improving executive function in children as young as 3 to 4 years of age (Dowsett & Livesey, 2000; Rueda, Rothbart, McCandliss, Saccomanno, & Posner, 2005; Tang, Yang, Leve, & Harold, 2012) through adulthood (Franklin & Franklin, 2012; Goudreau & Knight, 2018; Kramer, Larish, & Strayer, 1995; Parker & Boutelle, 2009). Specific interventions have also been designed for clinical populations, such as the Unstuck and On Target Program for children on the autism spectrum (Kenworthy et al., 2014) and training programs for children and adults with ADHD (Klingberg et al., 2005; White & Shah, 2006). Together, these studies emphasize the malleable nature of executive function, and suggest that individuals with 3q29del would benefit from targeted executive function interventions from an early age.

While the present study is the first detailed description of executive function in individuals with 3q29del, it is not without limitations. The average age of our study subjects is young (mean=14.50±8.26 years); as such, a majority of individuals have not reached the age at onset for SZ and psychotic disorders. Longitudinal follow up of study participants is needed to determine if executive function abilities predict later-onset phenotypes. Additionally, the small sample size of the present study rendered our analyses exploratory, and we were likely underpowered for some comparisons, particularly in the neuroimaging analysis. Ongoing work by our team includes executive function measurements in a larger sample of individuals with 3q29del; we will aim to replicate the results of the present study in that cohort. Finally, we were unable to assess the relative effect of race and ethnicity on executive function in the current study, as our sample was overwhelmingly white and non-Hispanic. Current and future recruitment efforts will aim to include more underrepresented minorities so that future studies ideally have a more representative study sample.

The present study is the first to describe details of executive function in individuals with 3q29del. We identified significant global deficits in executive function, which were consistent between males and females. We found that executive function was not correlated with IQ, but was significantly associated with SZ spectrum symptom severity, ADHD diagnosis, and neuropsychiatric and neurodevelopmental comorbidity; the BRIEF-2 ADHD Form accurately discriminated between individuals with 3q29del with and without ADHD. This study, coupled with previous work by our team, emphasizes the central nature of executive function to the 3q29del phenotype beyond general intellectual ability, and highlights the need for executive function evaluation and interventions for all individuals with 3q29del. The malleable nature of executive function means that early intervention in this population will likely yield substantial gains in abilities and improvements in long-term outcomes and ability to function independently.

## Data Availability

All data produced in the present study are available upon reasonable request to the authors.

## Acknowledgements

We gratefully acknowledge our study population, the 3q29 deletion community, for their participation and commitment to research.

## Financial Support

This work was supported by the National Institute of Mental Health grant R01 MH110701 (PI Mulle) and R01 MH118534 (MPI Schultz/Mulle).

## Ethical Standards

The authors assert that all procedures contributing to this work comply with the ethical standards of the relevant national and institutional committees on human experimentation and with the Helsinki Declaration of 1975, as revised in 2008.

## Competing Interests

CAS reports receiving royalties from Pearson Assessments for the Vineland-3. The remaining authors have no competing interests to disclose.

## Author Contributions

RMP performed analysis, created figures and tables, and wrote the manuscript. ES performed analysis, created figures and tables, wrote the manuscript, and collected data. KD and AEG performed analysis of neuroimaging data. KA, RE, RG, KG, and DMN collected data. CAS, CK, SP, JFC, and TLB evaluated study participants. SS led the collection of neuroimaging data and supervised analysis of neuroimaging data. EFW supervised data collection efforts. MMM planned and executed the study. JGM planned and executed the study and was the principal investigator responsible for study coordination.

## Notes

### Author Declarations

IRB of Emory University gave ethical approval for this work. IRB of Rutgers University gave ethical approval for this work.

