## Supplemental Information for "Beyond IQ: Executive function deficits and their relation to functional, clinical, and neuroimaging outcomes in 3q29 deletion syndrome"

**Supplemental Methods**

*Measures*

Executive function was assessed using the Behavior Rating Inventory of Executive Function, Second Edition (BRIEF-2), for participants 18 years of age and younger (n=26) or the Behavior Rating Inventory of Executive Function for Adults (BRIEF-A) for participants over 18 years of age (n=6) (Gioia, Isquith, Guy, & Kenworthy, 2015; Roth & Gioia, 2005). Study instruments were completed by the parent or guardian of the study subject via the publisher’s website (PARiconnect). The BRIEF-2 and BRIEF-A are questionnaire-based tools designed to assess executive function behaviors relevant to everyday life using a 3-point Likert scale. Items on the BRIEF-2 and BRIEF-A are grouped into nine distinct subscales that capture different components of executive function; a Global Executive Composite score that incorporates ratings from all subscales is also computed to reflect the subject’s overall level of executive function (Table S1). The BRIEF-2 and BRIEF-A are ecologically valid measures of the multidimensional nature of impairments in executive function, with high internal consistency and test-retest reliability (Gioia et al., 2015; Roth & Gioia, 2005). The BRIEF-2 and BRIEF-A are age- and sex-normed, with a mean of 50 and standard deviation of 10. Higher scores on the BRIEF-2 and BRIEF-A indicate more severe impairment in executive function abilities. The BRIEF-2 ADHD Form was used to compare to clinical ADHD diagnoses in our study population. The three validity scales of the BRIEF (negativity, infrequency, and inconsistency) were evaluated for each participant to ensure the reliability of responses; no participants had elevations of concern. General cognitive ability was evaluated using the Differential Ability Scales, Second Edition (DAS-II) (Elliott, Murray, & Pearson, 1990), for individuals under 18 years of age (n=24) or the Wechsler Abbreviated Scale of Intelligence, Second Edition (WASI-II) (Wechsler, 1999), for individuals 18 years of age and older (n=8). Adaptive behavior was assessed using the Vineland Adaptive Behavior Scales, Third Edition, Comprehensive Parent/Caregiver Form (Vineland-3) (Sparrow, Cicchetti, & Saulnier, 2016). Psychosis symptoms were assessed using the Structured Interview for Psychosis-Risk Syndromes (SIPS) for individuals 8 years of age and older (n=23) (Miller et al., 2003). Diagnoses of neurodevelopmental and neuropsychiatric phenotypes were reached using gold-standard evaluations and clinician best estimate diagnosis. Additional details regarding the assessments used in the present study have been previously described (Murphy et al., 2018; Sanchez Russo et al., 2021), and information about cognitive ability and adaptive behavior has been reported elsewhere (Klaiman et al., 2022; Sanchez Russo et al., 2021).

*Neuroimaging*

High-resolution structural MRI scans were performed on a subset of participants who had no contraindication (n=24) on a 3T Siemens Magnetom Prisma scanner at the Emory University Center for Systems Imaging Core using a 32-channel head coil and an 80mT/m gradient. T1-weighted 3D images were acquired in the sagittal plane using a single-echo MPRAGE sequence (Brant-Zawadzki, Gillan, & Nitz, 1992) with the following parameters: TE=2.24ms, TR=2400ms, TI=1000ms, bandwidth=210Hz/pixel, FOV=256x256mm, resolution=0.8mm isotropic. T2-weighted 3D images were acquired in the sagittal plane using a SPACE sequence (67) with the following parameters: TE=563ms, TR=3200ms, bandwidth=745Hz/pixel, FOV=256x256mm, resolution=0.8mm isotropic. Scans were processed with the HCP “minimal pre-processing” pipeline to remove spatial artifacts or distortions, align images to MNI space, and obtain brain masks/parcellations (Glasser et al., 2013). One subject was excluded from volumetric analyses due to motion artifacts but was retained in analyses involving radiological findings established by a board-certified neuroradiologist (Sefik et al., 2022). FreeSurfer software (https://surfer.nmr.mgh.harvard.edu) was used for automated segmentations, based on probabilistic information estimated from a manually labeled training set (Fischl et al., 2002). Using this approach, we extracted volumetric measures for cerebellar cortex and white matter to distinguish the two primary tissue types of the cerebellum. Total cerebellar volume was defined as the sum of these measures. Volumetric measures for subregions of the cerebellar cortex were extracted using the automatic cerebellum anatomical parcellation using U-Net with locally constrained optimization (ACAPULCO) (Han, Carass, He, & Prince, 2020) and subregional segmentation errors were manually corrected in 2D and 3D formats for increased anatomical accuracy using ITK-SNAP (http://www.itksnap.org/). Details on quality control have been previously reported (Sefik et al., 2022).

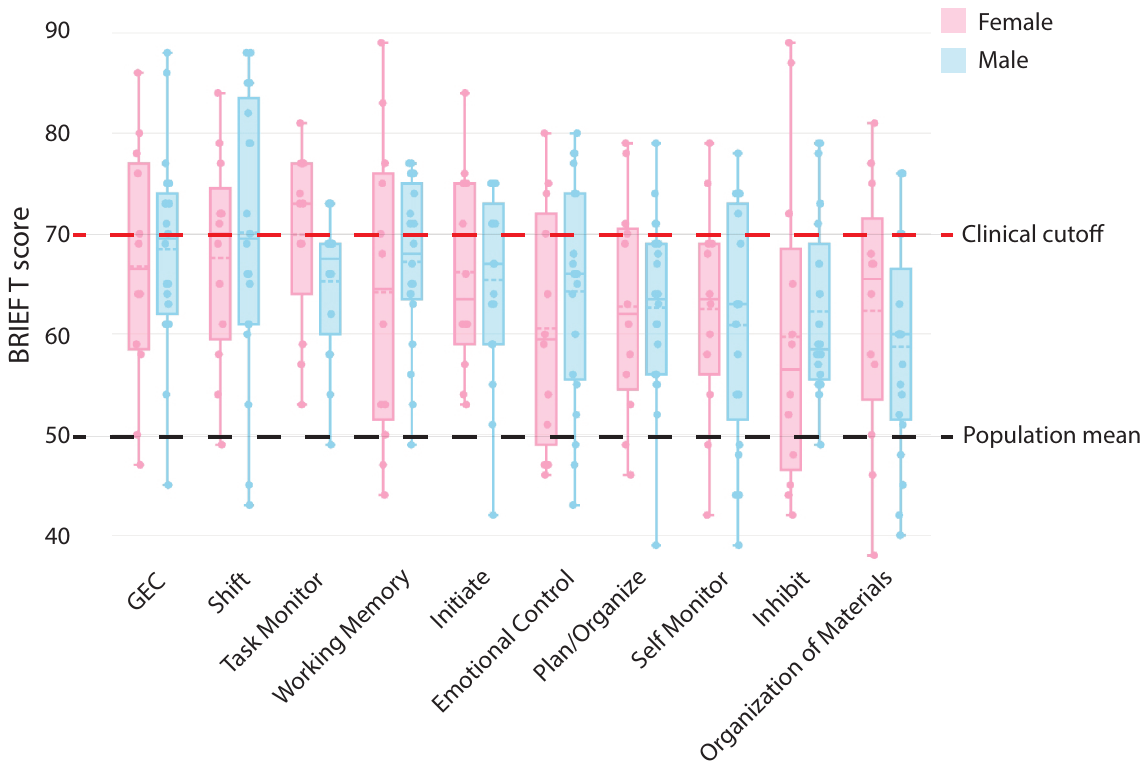

**Figure S1.** Distribution of T scores on the BRIEF GEC and subscales for male (n=20) and female (n=12) study participants with 3q29del.

3q29del, 3q29 deletion syndrome; BRIEF, Behavior Rating Inventory of Executive Function; GEC, Global Executive Composite

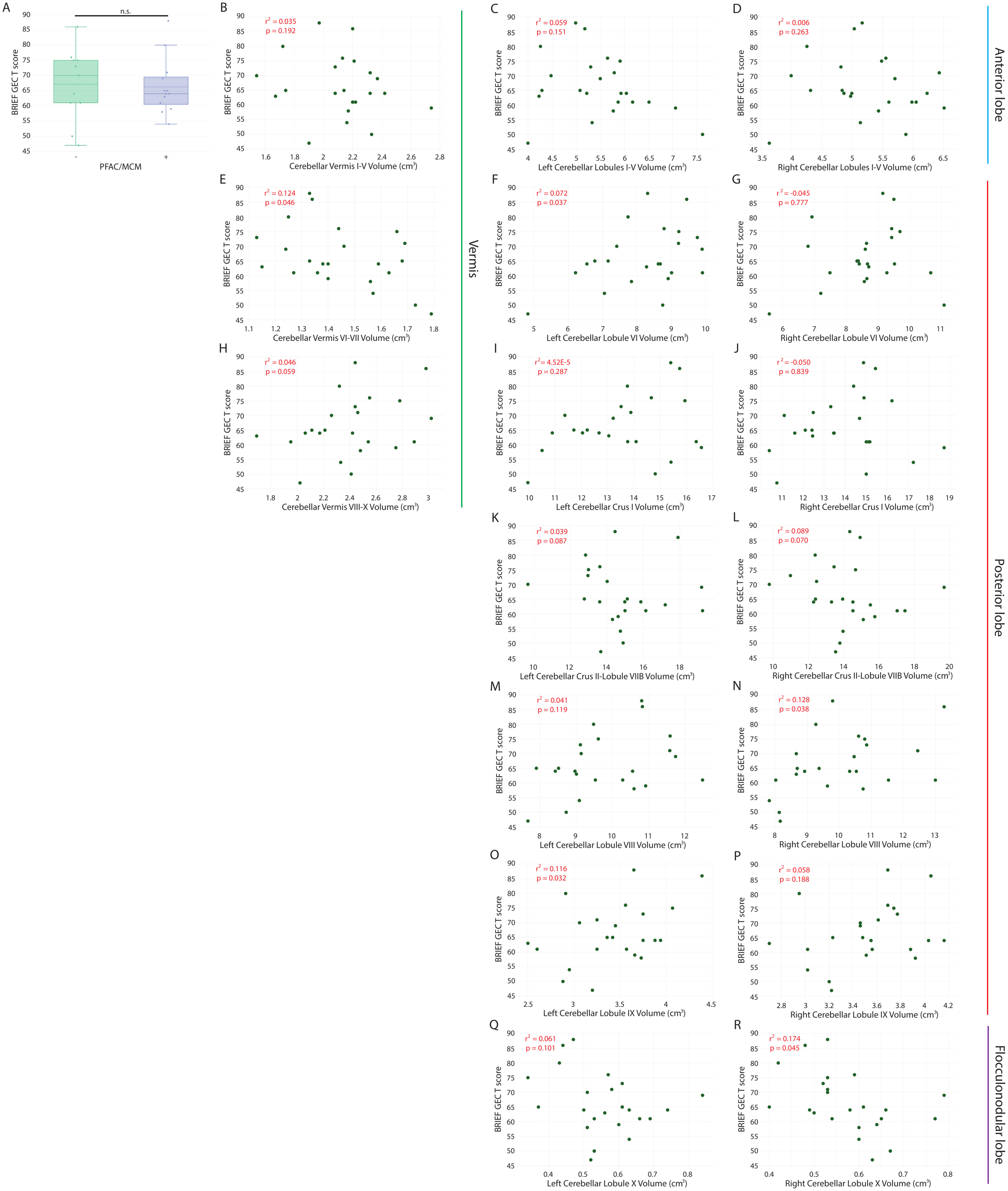

**Figure S2. A)** Distribution of T scores on the BRIEF GEC for study participants with 3q29del with (n=13) and without (n=11) posterior fossa arachnoid cyst/mega cisterna magna (PFAC/MCM). **B, E, H)** Correlation between BRIEF GEC T scores and volume of subregions of the vermis of the cerebellar cortex for study participants with 3q29del (n=23), divided into the **B)** cerebellar vermis I-V, **E)** cerebellar vermis VI-VII, and **H)** cerebellar vermis VIII-X. **C-D)** Correlation between BRIEF GEC T scores and volume of subregions of the anterior lobe of the cerebellar cortex for study participants with 3q29del (n=23), divided into the **C)** left hemisphere cerebellar lobules I-V and **D)** right hemisphere cerebellar lobules I-V. **F-G, I-P)** Correlation between BRIEF GEC T scores and volume of subregions of the posterior lobe of the cerebellar cortex for study participants with 3q29del (n=23), divided into the **(F)** left hemisphere cerebellar lobule VI, **(G)** right hemisphere cerebellar lobule VI, **(I)** left hemisphere cerebellar crus I, **(J)** right hemisphere cerebellar crus I, **(K)** left hemisphere cerebellar crus II-lobule VIIB, **(L)** right hemisphere cerebellar crus II-lobule VIIB, **(M)** left hemisphere cerebellar lobule VIII, **(N)** right hemisphere cerebellar lobule VIII, **(O)** left hemisphere cerebellar lobule IX, and **(P)** right hemisphere cerebellar lobule IX. **Q-R)** Correlation between BRIEF GEC T scores and volume of subregions of the flocculonodular lobe of the cerebellar cortex for study participants with 3q29del (n=23), divided into the **(Q)** left hemisphere cerebellar lobule X and **(R)** right hemisphere cerebellar lobule X.

3q29del, 3q29 deletion syndrome; BRIEF, Behavior Rating Inventory of Executive Function; GEC, Global Executive Composite

**Table S1.** BRIEF scale and subscale descriptions.

| **BRIEF scale** | | **Description** |
| --- | --- | --- |
| Global Executive Composite (GEC) | | Summary measure that represents and individual’s overall level of executive functioning |
| *Subscales* | |  |
|  | Shift | Ability to switch/alternate freely between one activity/situation/aspect of problem to another as circumstances demand; problem-solving flexibility |
|  | Task Monitor | Ability to keep track of one’s own performance during or shortly after finishing a task to ensure accuracy/appropriate attainment of a goal; identify and correct mistakes during tasks |
|  | Working Memory | Ability to actively hold information in mind for the purpose of completing a task or making the appropriate response |
|  | Initiate | Ability to begin a task/activity; independently generate ideas, responses, or problem-solving strategies |
|  | Emotional Control | Ability to modulate emotional responses appropriately to situational demand/context |
|  | Plan/Organize | Ability to anticipate future events; set goals; develop steps ahead of time to carry out a task; bring order to information, actions, or materials to achieve an objective |
|  | Self Monitor | Ability to keep track of/have awareness of the impact of one’s own behavior on other people/outcomes |
|  | Inhibit | Ability to control impulses; to appropriately stop behavior at the proper time/context |
|  | Organization of Materials | Ability to ensure orderliness of work, play, storage spaces (e.g., desks, lockers, bedrooms); organize/keep track of/clean up belongings |

**Table S2.** BRIEF scores for study participants with 3q29del (n = 32). P values are comparing 3q29del mean scores to the general population mean of 50 via one-sided, one-sample Student’s t tests. Subscales are reported in descending order of mean severity.

3q29del, 3q29 deletion syndrome; BRIEF, Behavior Rating Inventory of Executive Function; GEC, Global Executive Composite; SD, standard deviation

| **Scale** | **T score mean ± SD** | **Range** | **P value** | **Cohen’s d** |
| --- | --- | --- | --- | --- |
| GEC | 67.81 ± 10.68 | 45 - 88 | 6.33E-11 | 1.67 |
| Shift | 69.16 ± 12.52 | 43 - 88 | 4.45E-10 | 1.53 |
| Task Monitor | 67.00 ± 7.80 | 49 - 81 | 8.65E-14 | 2.18 |
| Working Memory | 66.06 ± 11.12 | 44 - 89 | 1.57E-09 | 1.44 |
| Initiate | 65.69 ± 9.29 | 42 - 84 | 4.72E-11 | 1.69 |
| Emotional Control | 62.88 ± 11.36 | 43 - 80 | 1.91E-07 | 1.13 |
| Plan/Organize | 62.69 ± 9.60 | 39 - 79 | 1.01E-08 | 1.32 |
| Self Monitor | 61.50 ± 11.24 | 39 - 79 | 1.12E-06 | 1.02 |
| Inhibit | 61.31 ± 11.92 | 42 - 89 | 3.75E-06 | 0.95 |
| Organization of Materials | 60.09 ± 11.65 | 38 - 81 | 1.43E-05 | 0.87 |

**Table S3.** Results of simple linear regressions assessing the relationship between BRIEF T scores and sex.

| Outcome variable: BRIEF GEC T score | | | |
| --- | --- | --- | --- |
| Predictors | b | CI (95%) | p value |
| Intercept | 66.75 | 60.37 - 73.13 | <2E-16 |
| Sex (M) | 1.70 | -6.37 - 9.77 | 0.683 |
| R2/Adjusted R2 | 0.006/-0.027 | |  |
| Wald test statistics | 0.185 on 1 & 30 DF | p value | 0.670 |
| Outcome variable: BRIEF Shift T score | | | |
| Predictors | b | CI (95%) | p value |
| Intercept | 67.58 | 60.12 - 75.05 | <2E-16 |
| Sex (M) | 2.52 | -6.92 - 11.96 | 0.564 |
| R2/Adjusted R2 | 0.010/-0.023 | |  |
| Wald test statistics | 0.296 on 1 & 30 DF | p value | 0.590 |
| Outcome variable: BRIEF Task Monitor T score | | | |
| Predictors | b | CI (95%) | p value |
| Intercept | 69.92 | 65.45 - 74.39 | <2E-16 |
| Sex (M) | -4.67 | -10.32 - 0.098 | 0.126 |
| R2/Adjusted R2 | 0.087/0.056 |  |  |
| Wald test statistics | 2.844 on 1 & 30 DF | p value | 0.102 |
| Outcome variable: BRIEF Working Memory T score | | | |
| Predictors | b | CI (95%) | p value |
| Intercept | 64.17 | 57.56 - 70.77 | 1.69E-15 |
| Sex (M) | 3.03 | -5.32 - 11.39 | 0.520 |
| R2/Adjusted R2 | 0.018/-0.015 | |  |
| Wald test statistics | 0.550 on 1 & 30 DF | p value | 0.464 |
| Outcome variable: BRIEF Initiate T score | | | |
| Predictors | b | CI (95%) | p value |
| Intercept | 66.17 | 60.60 - 71.73 | <2E-16 |
| Sex (M) | -0.77 | -7.80 - 6.27 | 0.828 |
| R2/Adjusted R2 | 0.002/-0.032 | |  |
| Wald test statistics | 0.050 on 1 & 30 DF | p value | 0.825 |
| Outcome variable: BRIEF Emotional Control T score | | | |
| Predictors | b | CI (95%) | p value |
| Intercept | 60.58 | 53.86 - 67.30 | <2E-16 |
| Sex (M) | 3.67 | -4.83 - 12.17 | 0.393 |
| R2/Adjusted R2 | 0.025/-0.007 | |  |
| Wald test statistics | 0.776 on 1 & 30 DF | p value | 0.385 |
| Outcome variable: BRIEF Plan/Organize T score | | | |
| Predictors | b | CI (95%) | p value |
| Intercept | 62.75 | 57.00 - 68.50 | <2E-16 |
| Sex (M) | -0.10 | -7.38 - 7.18 | 0.979 |
| R2/Adjusted R2 | 2.63E-5/-0.033 | |  |
| Wald test statistics | 0.001 on 1 & 30 DF | p value | 0.978 |
| Outcome variable: BRIEF Self Monitor T score | | | |
| Predictors | b | CI (95%) | p value |
| Intercept | 62.50 | 55.78 - 69.22 | <2E-16 |
| Sex (M) | -1.60 | -10.10 - 6.90 | 0.695 |
| R2/Adjusted R2 | 0.005/-0.028 | |  |
| Wald test statistics | 0.148 on 1 & 30 DF | p value | 0.703 |
| Outcome variable: BRIEF Inhibit T score | | | |
| Predictors | b | CI (95%) | p value |
| Intercept | 59.75 | 52.64 - 66.86 | 5.51E-14 |
| Sex (M) | 2.50 | -6.49 - 11.49 | 0.620 |
| R2/Adjusted R2 | 0.011/-0.022 | |  |
| Wald test statistics | 0.323 on 1 & 30 DF | p value | 0.574 |
| Outcome variable: BRIEF Organization of Materials T score | | | |
| Predictors | b | CI (95%) | p value |
| Intercept | 62.33 | 55.43 - 69.24 | <2E-16 |
| Sex (M) | -3.58 | -12.32 - 5.15 | 0.427 |
| R2/Adjusted R2 | 0.023/-0.010 | |  |
| Wald test statistics | 0.702 on 1 & 30 DF | p value | 0.409 |

**Table S4.** Results of simple linear regressions assessing the relationship between BRIEF GEC T scores and measures of cognitive ability.

| Outcome variable: Composite IQ | | | |
| --- | --- | --- | --- |
| Predictors | b | CI (95%) | p value |
| Intercept | 59.86 | 26.25 - 93.46 | 0.001 |
| BRIEF GEC T score | 0.19 | -0.30 - 0.68 | 0.424 |
| R2/Adjusted R2 | 0.021/-0.011 | |  |
| Wald test statistics | 0.657 on 1 & 30 DF | p value | 0.424 |
| Outcome variable: Nonverbal IQ | | | |
| Predictors | b | CI (95%) | p value |
| Intercept | 54.69 | 26.41 - 82.98 | 4.40E-4 |
| BRIEF GEC T score | 0.30 | -0.11 - 0.71 | 0.146 |
| R2/Adjusted R2 | 0.069/0.038 |  |  |
| Wald test statistics | 2.229 on 1 & 30 DF | p value | 0.146 |
| Outcome variable: Verbal IQ | | | |
| Predictors | b | CI (95%) | p value |
| Intercept | 81.92 | 34.32 - 129.52 | 0.001 |
| BRIEF GEC T score | -0.03 | -0.73 - 0.66 | 0.920 |
| R2/Adjusted R2 | 3.41E-4/-0.033 | |  |
| Wald test statistics | 0.010 on 1 & 30 DF | p value | 0.920 |

**Table S5.** Results of simple linear regressions assessing the relationship between BRIEF GEC T scores and measures of psychosis symptom severity on the SIPS.

| Outcome variable: SIPS positive symptom dimension | | | |
| --- | --- | --- | --- |
| Predictors | b | CI (95%) | p value |
| Intercept | -6.88 | -26.00 - 12.23 | 0.232 |
| BRIEF GEC T score | 0.206 | -0.07 - 0.49 | 0.031 |
| R2/Adjusted R2 | 0.100/0.057 |  |  |
| Wald test statistics | 2.338 on 1 & 21 DF | p value | 0.141 |
| Outcome variable: SIPS negative symptom dimension | | | |
| Predictors | b | CI (95%) | p value |
| Intercept | 0.73 | -16.99 - 18.44 | 0.933 |
| BRIEF GEC T score | 0.11 | -0.15 - 0.37 | 0.397 |
| R2/Adjusted R2 | 0.038/-0.008 | |  |
| Wald test statistics | 0.820 on 1 & 21 DF | p value | 0.375 |
| Outcome variable: SIPS disorganization symptom dimension | | | |
| Predictors | b | CI (95%) | p value |
| Intercept | -4.87 | -13.84 - 4.10 | 0.242 |
| BRIEF GEC T score | 0.15 | 0.02 - 0.28 | 0..022 |
| R2/Adjusted R2 | 0.219/0.182 |  |  |
| Wald test statistics | 5.89 on 1 & 21 DF | p value | 0.024 |

**Table S6.** Results of simple linear regressions assessing the relationship between BRIEF GEC T scores and neurodevelopmental and neuropsychiatric diagnoses.

| Outcome variable: ASD diagnosis | | | |
| --- | --- | --- | --- |
| Predictors | b | CI (95%) | p value |
| Intercept | 0.87 | -0.29 - 2.03 | 0.113 |
| BRIEF GEC T score | -0.01 | -0.02 - 0.01 | 0.340 |
| R2/Adjusted R2 | 0.025/-0.008 | |  |
| Wald test statistics | 0.769 on 1 & 30 DF | p value | 0.388 |
| Outcome variable: SZ prodrome/psychosis diagnosis | | | |
| Predictors | b | CI (95%) | p value |
| Intercept | -0.66 | -1.93 - 0.60 | 0.153 |
| BRIEF GEC T score | 0.01 | -0.004 - 0.03 | 0.057 |
| R2/Adjusted R2 | 0.110/0.067 |  |  |
| Wald test statistics | 2.587 on 1 & 21 DF | p value | 0.123 |
| Outcome variable: ADHD diagnosis | | | |
| Predictors | b | CI (95%) | p value |
| Intercept | -1.29 | -2.22 - -0.36 | 5.69E-5 |
| BRIEF GEC T score | 0.03 | 0.01 - 0.04 | 3.08E-8 |
| R2/Adjusted R2 | 0.375/0.354 |  |  |
| Wald test statistics | 18.01 on 1 & 30 DF | p value | 1.95E-4 |
| Outcome variable: Anxiety disorder diagnosis | | | |
| Predictors | b | CI (95%) | p value |
| Intercept | -0.29 | -1.46 - 0.87 | 0.549 |
| BRIEF GEC T score | 0.01 | -0.01 - 0.03 | 0.169 |
| R2/Adjusted R2 | 0.049/0.017 |  |  |
| Wald test statistics | 1.534 on 1 & 30 DF | p value | 0.225 |
| Outcome variable: ID diagnosis | | | |
| Predictors | b | CI (95%) | p value |
| Intercept | 0.48 | -0.68 - 1.63 | 0.342 |
| BRIEF GEC T score | -0.002 | -0.02 - 0.01 | 0.783 |
| R2/Adjusted R2 | 0.002/-0.031 | |  |
| Wald test statistics | 0.057 on 1 & 30 DF | p value | 0.814 |
| Outcome variable: Graphomotor weakness diagnosis | | | |
| Predictors | b | CI (95%) | p value |
| Intercept | 0.37 | -0.63 - 1.36 | 0.419 |
| BRIEF GEC T score | 0.01 | -0.01 - 0.02 | 0.325 |
| R2/Adjusted R2 | 0.024/-0.008 | |  |
| Wald test statistics | 0.748 on 1 & 30 DF | p value | 0.394 |
| Outcome variable: Enuresis diagnosis | | | |
| Predictors | b | CI (95%) | p value |
| Intercept | 0.19 | -0.81 - 1.20 | 0.596 |
| BRIEF GEC T score | 0.0004 | -0.01 - 0.02 | 0.944 |
| R2/Adjusted R2 | 8.91E-5/-0.033 | |  |
| Wald test statistics | 0.003 on 1 & 30 DF | p value | 0.959 |
| Outcome variable: Cumulative neurodevelopmental/neuropsychiatric diagnoses | | | |
| Predictors | b | CI (95%) | p value |
| Intercept | -0.13 | -3..16 - 2.90 | 0.926 |
| BRIEF GEC T score | 0.05 | 0.002 - 0.90 | 0.039 |
| R2/Adjusted R2 | 0.130/0.101 |  |  |
| Wald test statistics | 4.481 on 1 & 30 DF | p value | 0.043 |

**Table S7.** Results of multiple linear regressions assessing the relationship between BRIEF GEC T scores and cerebellar regional and subregional volumes.

[See attached file]
